# Disentangling network topology and pathogen spread

**DOI:** 10.1101/2021.05.24.21257706

**Authors:** María Pérez-Ortiz, Petru Manescu, Fabio Caccioli, Delmiro Fernández-Reyes, Parashkev Nachev, John Shawe-Taylor

## Abstract

How do we best constrain social interactions to prevent the transmission of communicable respiratory diseases? Indiscriminate suppression, the currently accepted answer, is both unsustainable long term and implausibly presupposes all interactions to carry equal weight. Transmission within a social network is determined by the topology of its graphical structure, of which the number of interactions is only one aspect. Here we deploy large-scale numerical simulations to quantify the impact on pathogen transmission of a set of topological features covering the parameter space of realistic possibility. We first test through a series of stochastic simulations the differences in the spread of disease on several classes of network geometry (including highly skewed networks and small world). We then aim to characterise the spread based on the characteristics of the network topology using regression analysis, highlighting some of the network metrics that influence the spread the most. For this, we build a dataset composed of more than 9000 social networks and 30 topological network metrics. We find that pathogen spread is optimally reduced by limiting specific kinds of social contact – unfamiliar and long range – rather than their global number. Our results compel a revaluation of social interventions in communicable diseases, and the optimal approach to crafting them.

## 1 Introduction

Many diseases spread via close physical interactions. The interpersonal contact patterns that underlie disease transmission naturally form a network, where links join individuals that interact and disease spreads along these links. Computational models of infectious disease transmission dynamics can provide scenario analysis during epidemic outbreaks necessary to devise the effectiveness of public health interventions such as quarantine and vaccination. All such epidemiological models make assumptions about the underlying network of interactions, often without explicitly stating them. Contact network models, however, mathematically formalize this intuitive concept so that epidemiological calculations can explicitly consider complex patterns of interactions [2].

Contact patterns among hosts are considered as one of the most critical factors contributing to unequal pathogen transmission. Consequently, the field of social networks have proven very useful in modeling infectious diseases. Contact rates for example often vary because of differences in individual traits, including individual behavior, as well as changes in the overall contact patterns along time and across space [9]. However, these sources of heterogeneity are currently often not being considered for devising policies at country level. This is because the predominant SIR models fail to take into account heterogeneous human behaviours that shape a country’s social network and cannot intervene on the contagion dynamics with precision [24]. Even more, previous work has argued that models that assume an homogeneous and mixed population can only lead to one type of intervention [24]. This is, interventions that indifferently concern large subsets of the population or even the overall population (e.g. national lockdowns). Recent studies looking at the most effective post lockdown interventions have also shown that these depend strongly on the underlying contact network [5, 17, 4].

Models including explicit representations of network topologies have been advocated as a necessary improvement of classical epidemiological models at least since early 2000 (see, for instance, [20]). However, models’ variants supporting policy decision-making in the current crisis continue to ignore this stream of the literature; and the consequences of not properly considering social networks for intervention are not systematically discussed [24]. We aim to partly fill this gap through focusing on contact network epidemiology [25, 13, 10], simulating several sources of diversity of the contact patterns that underlie disease transmission and showing their effects on the spread.

Recent works have shown that networks with equal number of nodes and edges, but different network structure (e.g. different path lengths and clustering) lead to different infection curves [4]. However, most of the theoretical literature [25, 33, 26, 1, 30, 18] focuses on the effect of individual network properties (e.g. the degree distribution, or assortative mixing, or clustering), which are typically analyzed by means of controlled numerical experiments or analytical calculations where only the property of interest is varied while keeping the rest fixed. In a realistic context, however, altering the structure of a network means simultaneously changing different network metrics. This is because when one perturbs a network, e.g. to increase the variance of the degree distribution, many other network metrics can change at the same time, and it is difficult to disentangle the effect of different network metrics on the number of infections. Even more, the size and complexity of the space of possible network characteristics makes the derivation of optimal metrics of spread from empirical data infeasible, for any candidate model is bound to be underdetermined by the scale and fidelity of available data. Rather we need large-scale numerical simulations spanning the full horizon of empirically plausible network parameters within which any real world network is bound to lie. Here we simulate more than 9000 social networks and compute 30 network metrics, using univariate and multivariate analysis to find the most predictive metrics of the spread.

### Paper layout

The next subsection presents a summary of the methodology used and the main research findings. Section 2 then introduces the methodology and simulation design choices in full detail. Section 3 presents all our results and finally, Section 4 discusses the implications of our findings and outlines some conclusions.

### 1.1 Summary of methodology and research findings

#### Method

We apply the standard compartmental SEIR model to stochastic networks. To do so, we consider a graph representing individuals (nodes) and their interactions (edges). At a given time, an individual makes contact with a subset of random individuals from their set of close contacts (denominated local interaction) with certain probability and with a subset of individuals outside of their network (called global interactions) with a different probability. More specifically, in our model (inspired from [29]), we consider two different types of social interactions: i) *Local interactions*, i.e. with close contacts – individuals with whom one has non-cursory (e.g., repeated, sustained, and/or physical) interactions on a regular basis, such as housemates, family members, close coworkers, close friends, etc. and ii) *Global interactions*, i.e. with casual contacts – individuals with whom one has incidental, brief, or superficial contact on an infrequent basis (e.g., at the grocery store, on transit, at a public event, in the elevator). Global interactions are represented in the models in the form of a parallel mode of mean-field global transmission. We consider three well-established classes of social networks representative of a variety of animal and human social networks [31]: i) random homogeneous networks, also known as Erdos-Renyi, ii) heavy tailed networks or scale-free and iii) small world networks. These structures partially include aspects commonly observed in human interactions such as heterogeneity, heavy-tailed and broad degree distribution, transitivity, assortativity and community structure. Additionally, we employ a community-based graph generator [14] to construct a more diverse set of topologies covering a large range of the aforementioned parameters. For each graph, we compute 30 different topological network metrics, which we categorise in global and local metrics depending on whether they are computed only on the basis of nodes’ neighbours (local), or using the whole graph (global).

### 1.1.1 The effect of network connectivity

Our extensive simulations (>100 000 runs) show that diverse families of networks with the same average degree – the conventional measure of transmission and the often assumed decisive characteristic of a network – can vary widely in their disease burden (Figure 1.1.1). Specifically, we show that even for a fixed average degree the spread can change drastically, i.e. from 10% to more than 60% infected for average degree of 4, and from 30% to more than 80% for average degree of 20. This difference is particularly significant when the average degree is low (e.g. degree of 4). Even though the density function in both plots depends on the distribution of networks that we have generated in our experiments, it is noteworthy that there is great difference in the spread for networks with the same average degree. Despite this difference in the spread for networks of the same average degree, most works that aim to study network metrics that are predictive of the spread, or that estimate such metrics from real world datasets, focus on the degree distribution [28].

**Figure 1:**
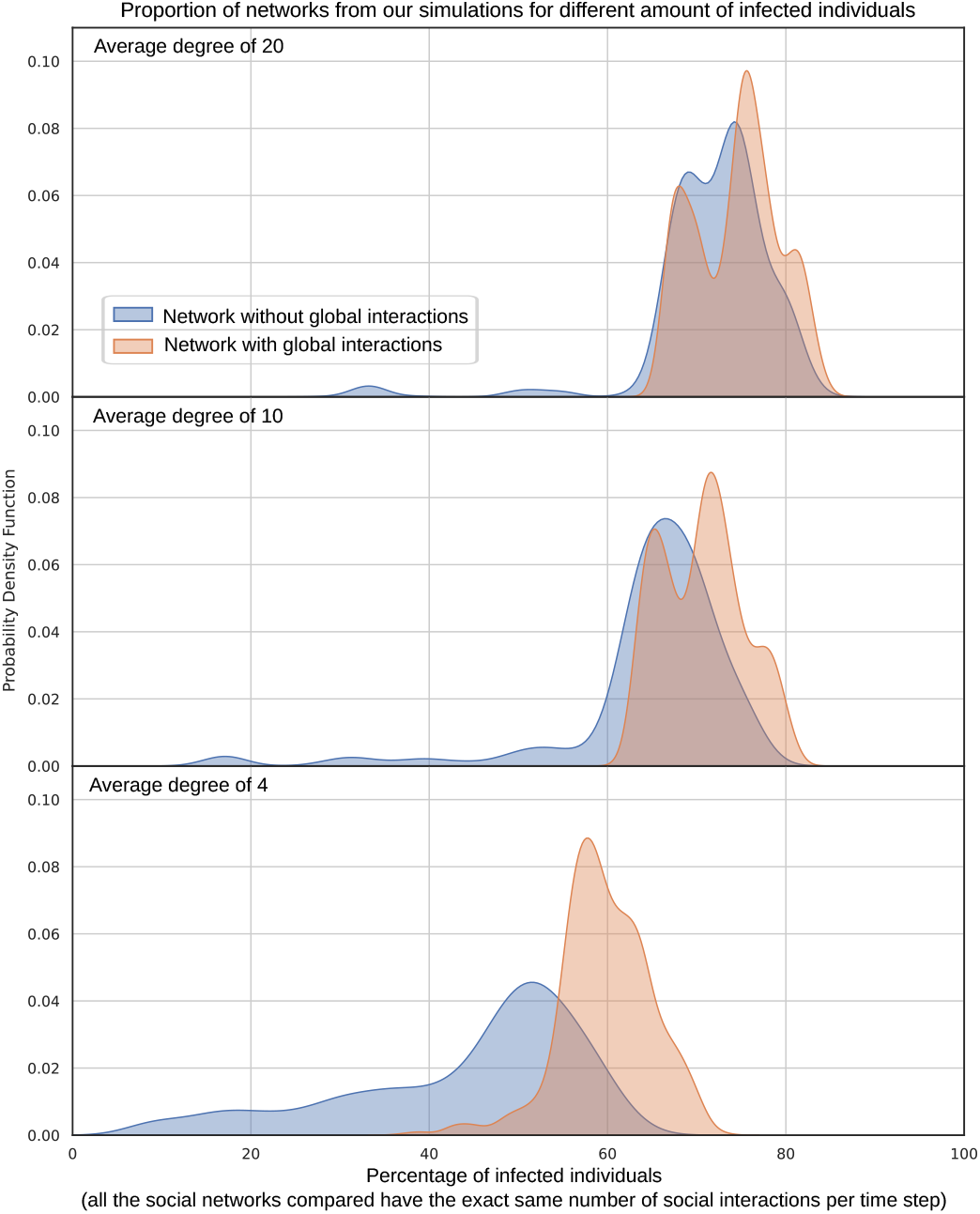
Proportion of networks from our simulations that result in a certain percentage of infected individuals, grouping contact networks by average degree and presence of global connections. In the simulation without global interactions individuals exclusively interact with their contact network, as opposed to the simulation with global interactions where individuals at each time step have a 20% chance of interacting with a random individual in the whole population and a 80% chance of meeting someone in their contact network. Importantly, for all the compared networks each individual on average only meets one other individual per time step, so independently of the average degree and globality all networks in this plot entail exactly the same number of social interactions per time step.

The second major insight is that not all human interactions affect the spread equally [35, 6]. Figure 1.1.1 shows the difference between i) weakly and strongly connected networks (as represented by the average degree) and ii) networks with/without global interactions. The spread increases for all strongly and globally connected networks, even when the amount of social interactions per time step is kept constant for all of these simulations. Again, global interactions affect specially the results of low connectivity networks (degree of 4). This striking variation reflects a neglected truth – social contacts differ in their effect on spread. Networks with the same volume of interaction per time step yield different levels of disease burden dependent on the form, not just the volume of observed connectivity. Critical here is the range of an interaction: the more local it is, the less the impact on overall spread [6].

In summary this first set of experiments show that obviating the underlying contact network through which disease spreads greatly impacts disease spread projections and consequently the potential usefulness of social interventions. Such a comprehensive characterisation of the impact on spread of each specific aspect of a network would allow to identify social interventions with optimal efficiency. Optimality is here formally defined by the marginal reduction in disease burden per ablated contact. The experiment additionally shows that pathogen spread is optimally reduced by limiting specific kinds of social contact – unfamiliar and long range – rather than their global number.

### 1.1.2 The effect of network families

Figure 1.1.2 and 1.1.2 show the results in percentage of infections (total and peak) for different well-known families of networks: i) Erdos Renyi, ii) scale-free and iii) small world. We can see that Erdos Renyi networks, the assumption made often in epidemiological models, overestimate the infections when compared to networks that resemble real-world ones better. Scale-free often leads to less infections than Erdos Renyi but a higher peak of infection. Small world networks are the most resilient to infections, especially for societies without global interactions. This shows that there exist network geometries in which contact does not have to be minimised in order to reduce the spread of the pathogen and that at fixed average degree there are topologies that are better than others. These cases are worth studying.

**Figure 2:**
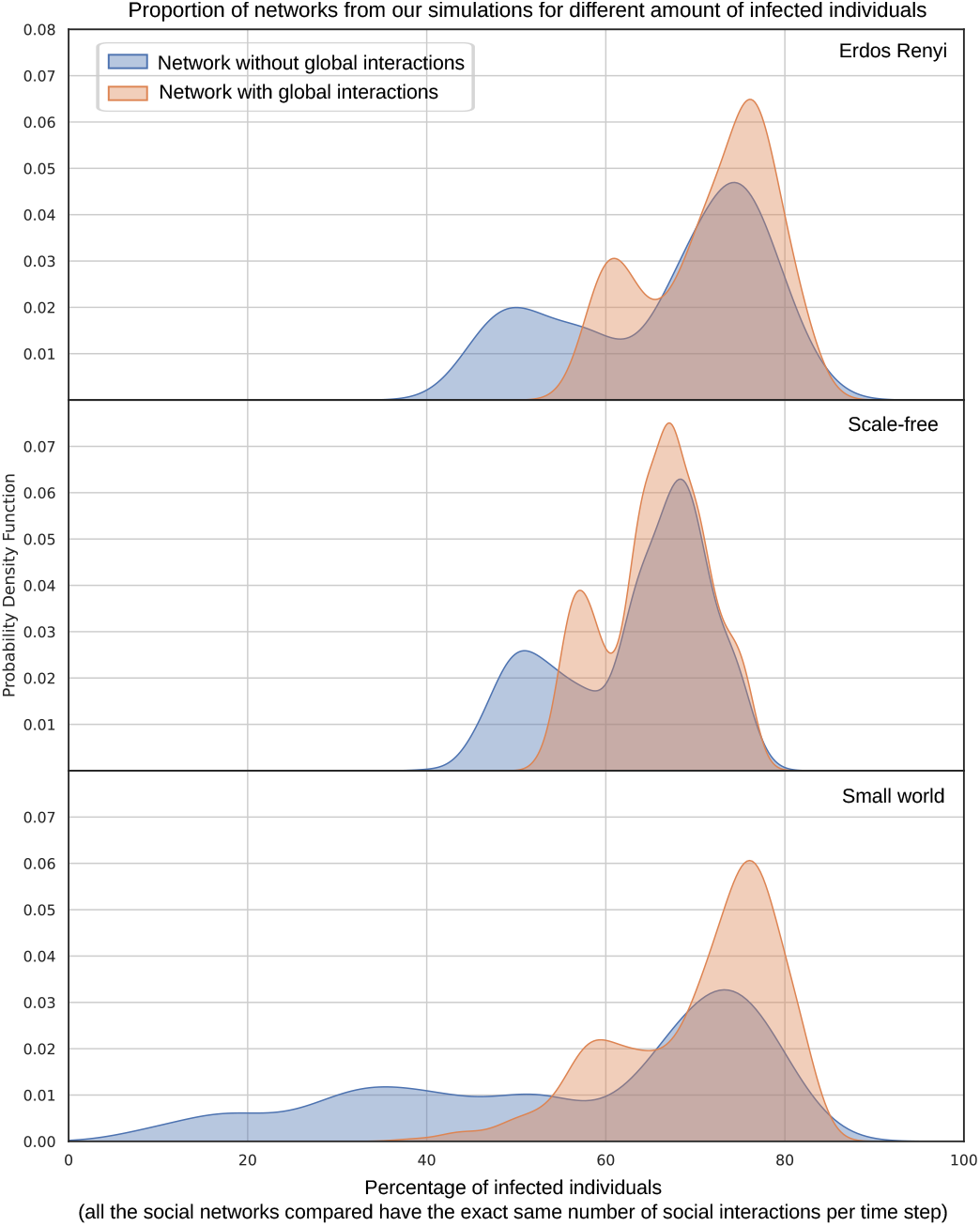
Proportion of networks from our simulations that result in a certain percentage of infected individuals, grouping contact networks by well-known families.

### 1.1.3 The effect of network metrics

Figure 1.1.3 shows a scatter plot of the relationship between several network metrics and the cumulative percentage of infected. Both these plots and the correlation analysis in the result section show that novel topology metrics (e.g. global efficiency and algebraic connectivity) show a stronger relationship than metrics proposed before, such as metrics of the degree distribution or the spectral radius. This motivates the need for computing such metrics from contact tracing, using mobility data that is already available or simply through social studies for different populations.

**Figure 3:**
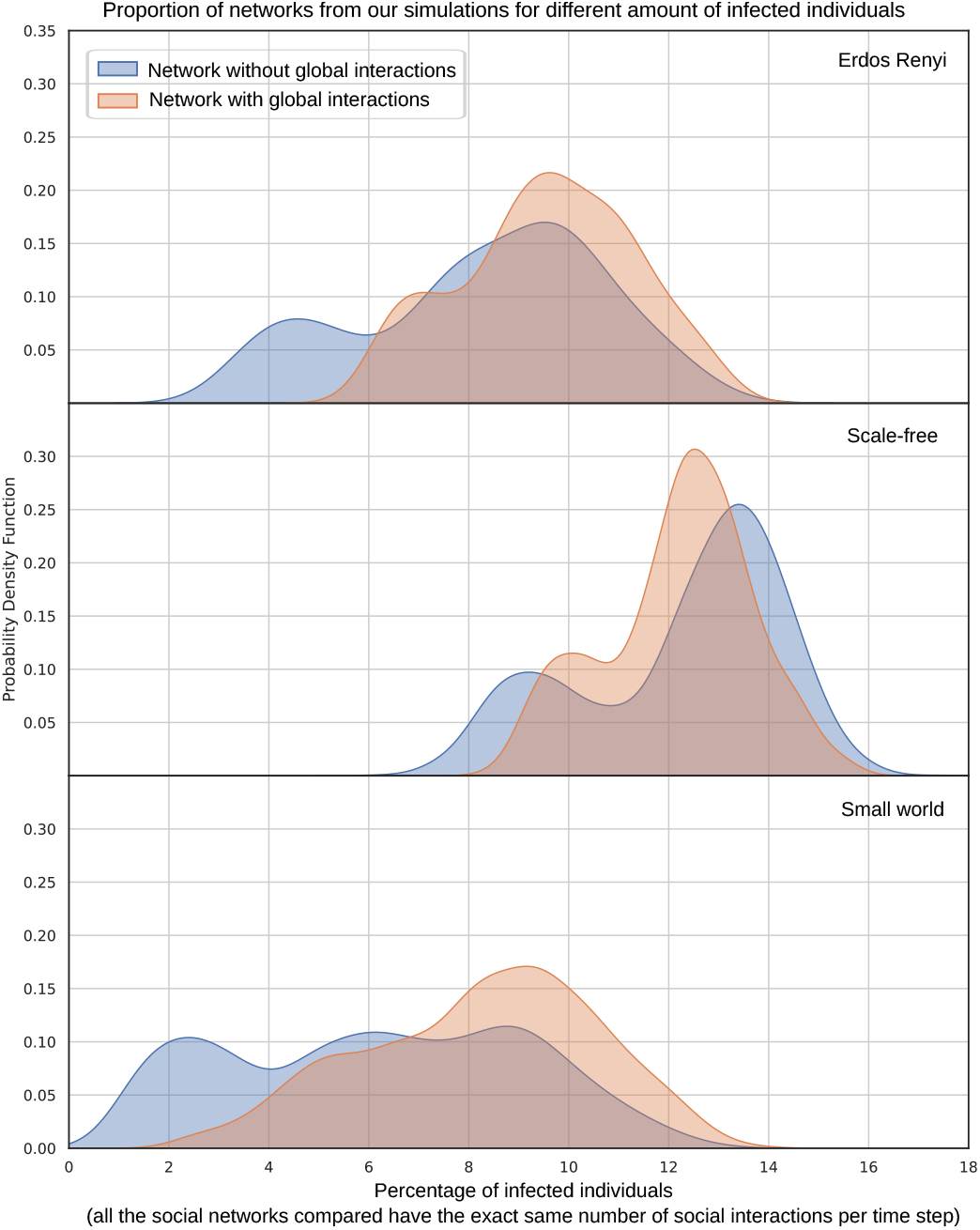
Proportion of networks from our simulations that result in a certain peak of infected individuals, grouping contact networks by well-known families.

**Figure 4:**
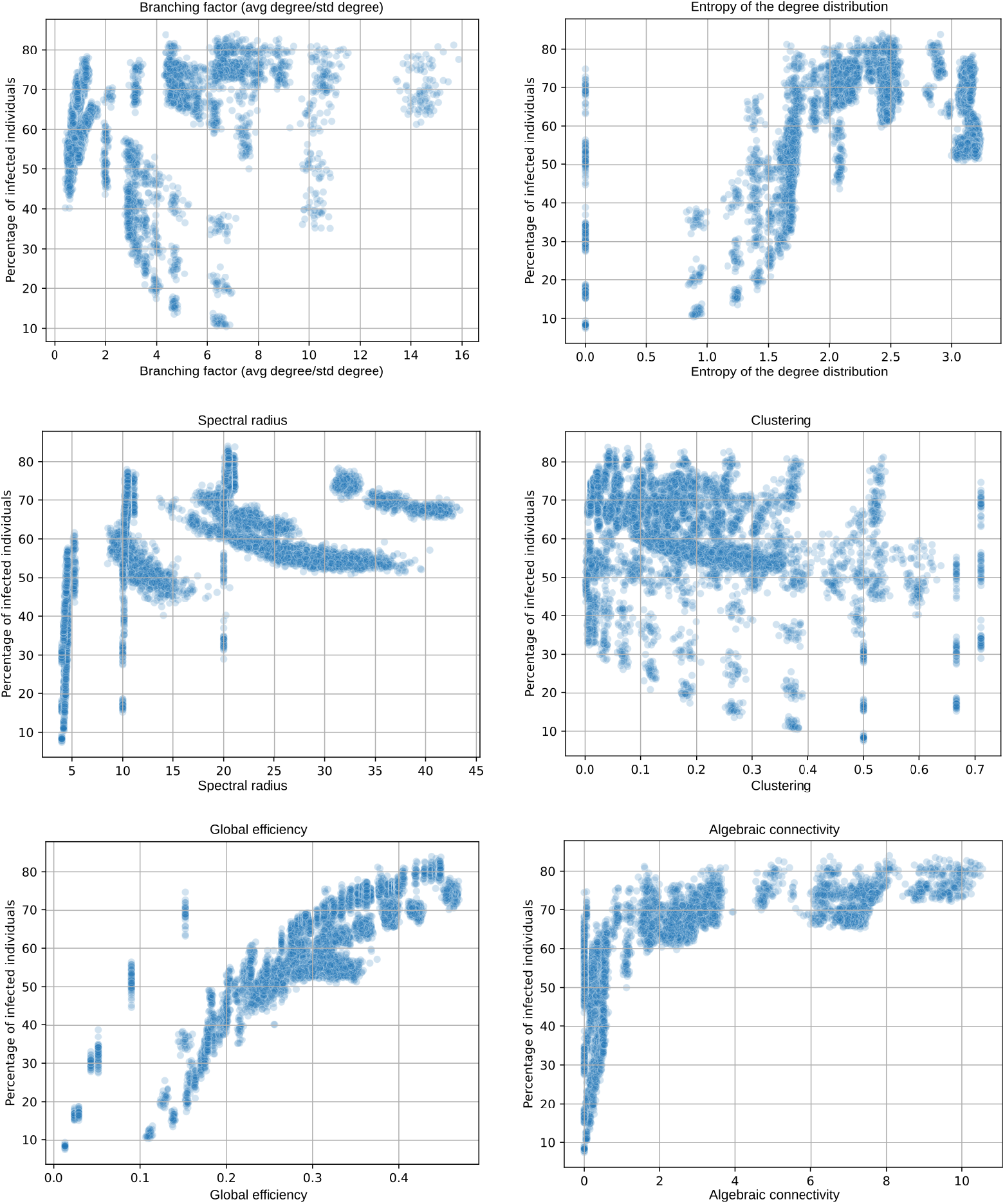
Scatter plots of a selection network metrics vs the cumulative percentage of infected for all the networks (also including community-based networks).

**Figure 5:**
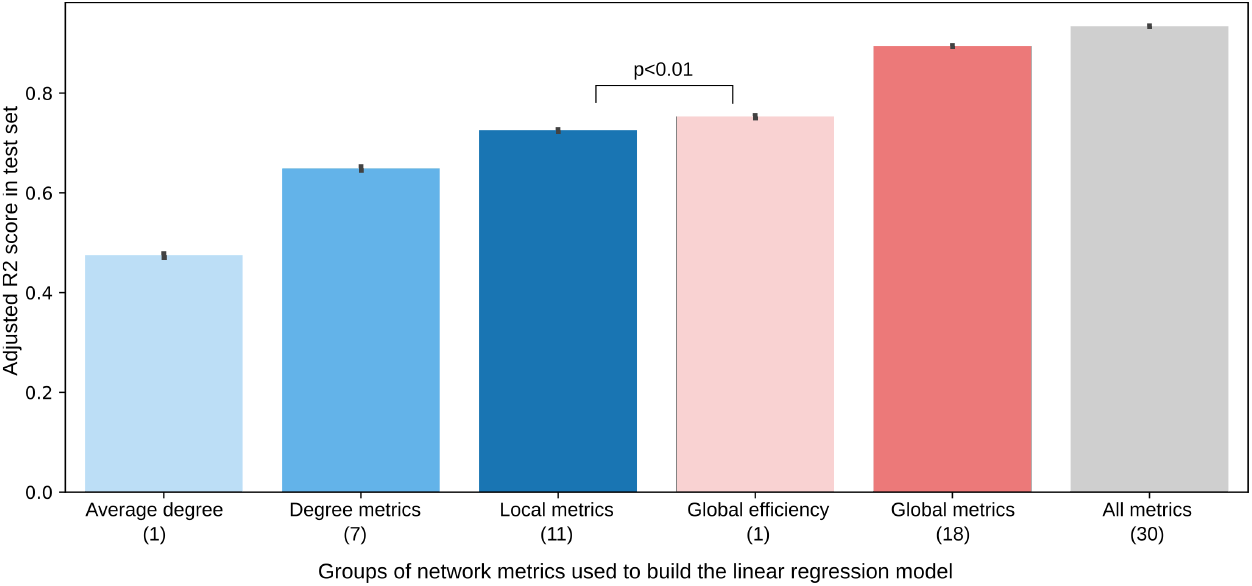
Bar chart of the test set performance (y-axis, measured as adjusted R2 score) of the different regression models built on groups of features (x-axis, each group including in parenthesis the total number of features). The chart includes the confidence intervals computed over 100 random data holdouts (80% train and 20% test). All comparisons are statistically significant for p<0.01.

We investigated the influence of metrics on the pathogen spread with a multivariate linear regression analysis. The results are shown in Figure 1.1.3. We report adjusted *R*^2^ computed on the test set of 100 random holdouts with 80% of data for training and 20% for testing. The first insight is that a combination of network metrics can accurately predict the variability of the spread. When considering all metrics, the linear regression model obtains an adjusted *R*^2^ of 0.934 (in comparison to a nonlinear one that obtains 0.987). The small confidence intervals suggest highly stable results. This figure also shows the predictive power of different groups of metrics. First, we can see that average degree alone is not enough to predict the spread accurately. Considering additional degree metrics (skewness, entropy, etc.) improves the predictive performance (*R*^2^ from 0.473 to 0.631), further improved when taking into account the remaining local metrics (*R*^2^ from 0.631 to 0.710). However, the model built using only global efficiency achieves better performance than the model that uses all degree metrics and the model that uses all local metrics (*R*^2^ of 0.747). However, the data has strong multicollinearity, precluding us from drawing further conclusions from the coefficients of these models. Interestingly, both degree distribution and local metrics are the metrics most often collected in studies and considered in epidemiological simulations. Additional global metrics complement global efficiency and increase the performance further. A non-linear regression model trained with global metrics alone was able to accurately predict the spread as well as a model trained with all metrics. We have shown a wide set of scenarios where outcomes can be reliably predicted from a model built using the topological properties of the underlying social network. We show this to be true across a full range of empirically-informed plausible network configurations. These results, of course, point to the question of what are the network metrics that represent the real world. Although some studies and datasets aimed at this exist [28], these works often focus exclusively on metrics such as the average of the degree distribution. Instead, we have shown that we urgently need reliable estimators of other crucial network metrics.

The univariate correlation analysis confirms that global efficiency, algebraic connectivity and the average closeness of a network have the highest influence on the spread. All of these metrics relate to the path lengths between nodes and global connectivity of the social graph. Both contact tracing or mobility data that are already being recorded can be used for example to infer path lengths in different societies [27, 21, 22] and thus restrict the social graph of simulations used in policy making.

## 2 SEIR on a contact network

We begin with an overview of compartmental models, the traditional approach to modeling infectious disease dynamics. We then introduce contact and social network epidemiology, which model the spread of infectious disease through heterogeneous populations. These methods, coupled with powerful computational methods, can help to address public health challenges and optimize epidemic control strategies.

We then describe the model used for the simulations, which is an extension of the widely-used compartmental SEIR model to social networks. We further review some of the characteristics of human interaction networks and describe the families of networks considered in this paper. Finally, we include a list of all the network metrics computed in this work for the analysis of the most influential variables for the spread.

### 2.1 Compartmental modelling

Compartmental models subdivide host populations by disease status. The adjective compartmental comes from viewing the disease states as compartments into and out of which individuals move throughout the epidemic. This is the case of a simple and widely used model named SIR, which tracks the movement of hosts among three states: (1) susceptible (S), meaning that the individual has never had the disease and is susceptible to being infected; (2) infected (I), meaning that the individual currently has the disease and can infect other people; and (3) resistant (R), meaning that the individual does not have the disease, cannot infect others, and cannot be infected.

The model then evolves in discrete time steps [12]: (1) Each susceptible individual draws a uniformly random person from the population. If the person drawn is infected, then the susceptible individual changes his state to infected with probability *β*. (2) Each infected individual changes his state to resistant with probability *ν*. (3) Each resistant individual remains resistant. This model makes several important assumptions, e.g. infected hosts are assumed to have contacts with random individuals from the population according to a Poisson process that yields an average contact rate of *β* per unit time. Disease transmission then occurs if and only if the individual at the receiving end of the contact is susceptible. Infectious hosts leave the infectious state at an average rate *ν* either by recovering and becoming immune or by dying. In the limit of a large host population, this process can be modeled by the following coupled nonlinear differential equations:

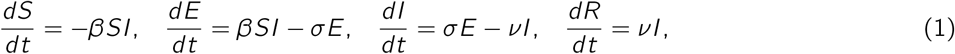

where *S*(*t*), *I*(*t*), *E*(*t*) and *R*(*t*) are the numbers of susceptible, infected, exposed and recovered hosts, respectively. The model ignores the birth and death of susceptibles, the total population size *N* is static.

Although compartmental SIR models have proven to be quite useful in modeling epidemics, they often do not properly model some important aspects of disease spread, since they assume a fully mixed, homogeneous population which may not adequately reflect reality.

### 2.2 Contact network epidemiology

Many diseases spread through human populations via close physical interactions. The interpersonal contact patterns that underlie disease transmission can naturally be thought to form a network, where links join individuals who interact with each other. During an outbreak, disease then spreads along these links. All epidemiological models make assumptions about the underlying network of interactions, often without explicitly stating them. Contact network models, however, mathematically formalize this intuitive concept so that epidemiological calculations can explicitly consider complex patterns of interactions [2].

In a contact network, each person translates into a vertex, and contacts among people translate into edges that connect appropriate vertices [25]. The number of edges emanating from a vertex is called the degree of the vertex and indicates the number of possible contacts that can lead to disease transmission to or from an individual.

### 2.3 Network SEIR Model

We apply the standard compartmental SEIR model to stochastic networks. To do so, we consider a graph *G* representing individuals (nodes) and their interactions (edges). Each node individual *i* in the graph has associated a current state: state *X*_*i*_ can be *S* (susceptible), *E* (exposed), *I* (infected) or *R* (recovered). At a given time, an individual *i* makes contact with a subset of random individuals from their set of close contacts (denominated local interaction) with probability *p*_*l*_ and with a subset of individuals outside of their network (called global interactions) with probability *p*_*g*_. More specifically, in our model (inspired from [29]), we consider two different types of contacts:

- **Close contacts** – individuals with whom one has non-cursory (e.g., repeated, sustained, and/or physical) interactions on a regular basis, such as housemates, family members, close coworkers, close friends, etc. This set of close contacts the population is defined through the contact network.
- **Casual contacts** – individuals with whom one has incidental, brief, or superficial contact on an infrequent basis (e.g., at the grocery store, on transit, at a public event, in the elevator) – are also represented in these models in the form of a parallel mode of mean-field global transmission.

When a susceptible individual interacts with an infectious individual they become exposed with probability *β* and transition towards infected with rate of progression *σ*. The model takes three pathogen parameters: probability of transmission *β* given contact, rate of progression *σ* and rate of recovery *γ*. These have been initialised to estimated parameters for SARS-CoV-2.

The probability of transitioning from exposed to infected and from infected to recovered remain the same than in the standard SEIR model (described in more detail in Section 2). However, the probability that a susceptible individual *i* moves to the exposed state needs to be specified by the contact network of the individual. We define it as:

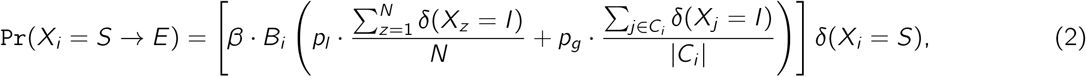

where *δ* is an indicator function such that *δ*(*X*_*i*_ = *A*) = 1 if the state of *X*_*i*_ is A, or 0 if not, *C*_*i*_ denotes the set of close contacts of node i and *B*_*i*_ is a factor accounting for the budget of interactions of individual *i* to its contact network at each time step. The terms 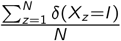 and 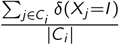 thus correspond respectively to the percentage of infected individuals in the population and in the contact network of individual *i*, respectively. The product of the network locality parameters (*p*_*l*_ and *p*_*g*_) and the transmission parameter set the weight of transmission among close (local) and casual (global) contacts in the modeled population.

If *B*_*i*_ is set to 1, then individual *i* makes contact with exactly 1 individual each time step of the simulation (either in the contact network or outside of it with probabilities *p*_*l*_ and *p*_*g*_). However, this makes the assumption that individuals highly connected have the same interactions per time step than individuals with a small contact network. In other words, it assumes an equal budget of interactions per individual, which may not be realistic in the case of super-spreaders. In fact, previous work has shown a super-linear association between the number of contacts and their duration [8], indicating the possibility that super-spreaders need to be defined not only in number of connections but also in intensity. This is, the more distinct interactions one individual has, the larger is the average time dedicated to those interactions.

We set to compare both assumptions regarding this budget of human interactions in our simulations. However, as it is obvious, we can not compare the result of two simulations with different total of human interactions, as the simulation with larger number of interactions would most probably lead to a larger spread of the pathogen. To appropriately compare the spread for an equal and unequal budget of interactions, we set to maintain the total number of interactions across the population per time step. This is, we first test *B*_*i*_ = 1 ∀*i*, where all individuals are assumed to be mixed similarly. We then compare these results to the case of 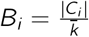, where 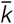 is the average degree for the population. This sets a linear relationship between *B*_*i*_ and |*C*_*i*_|, with individuals with larger contact networks having a larger interaction budget, while still maintaining the total number of interactions for the whole population. We additionally experiment with different *p*_*l*_ and *p*_*g*_, to see the difference that local/global interactions have on the spread.

Our proposed SEIR model on a social network has some limitations that we do not explore in this paper. For example, the amount of local and global interactions would often differ among individuals, with super-spreaders probably having a higher probability of global/random interactions. Instead of proposing Eq. (2) as a realistic model for the pathogen transmission, we aim to show the difference that certain choices of the model can have in the simulation results.

### 2.4 Characteristics of human interaction networks

There are some properties that are shared by human interaction networks. We review some of these now:

- **Heterogeneity** The degree (size of the contact network per individual) varies across individuals and groups of like-individuals (e.g., age groups). Groups of individuals may differ in the numbers of within- and between-group contacts they make.
- **Broad degree distribution** Most individuals have roughly average connectivity (degree), but there is individual variation around the mean degree (this is in contrast with, scale-free networks where most individuals have very low degree and the mode is often well below the mean).
- **Heavy-tailed degree distribution** A small number of individuals have many more contacts than average, so the degree distribution tends to have a relatively long right tail.
- **Assortativity** There tends to be correlation in degree between adjacent nodes in the contact network. That is, highly-connected individuals tend to have highly-connected contacts.
- **Transitivity (or clustering)** Individuals A and B are relatively likely to be contacts of each other if they both share a mutual contact C.
- **Community Structure** Contact networks often have communities of individuals (groups of nodes) that are more likely to be contacts of each other than they are to be with individuals from another community.

### 2.5 Families of networks considered

We consider three classes of social networks: i) *random homogeneous networks*, also known as *Erdos-Renyi*, ii) *heavy tailed networks or scale-free* and iii) *small world networks*. Part of these networks have been shown to be the representative of a variety of animal and human social networks [31] and cover some of the characteristics outlined:

- **Random homogeneous networks** are the underlying assumption of most compartmental models. The degree distribution can be approximated by a Poisson and peaks around the average, thus denoting a statistical homogeneity of the nodes. These networks are governed only by stochasticity and thus do not represent any structural properties that one would expect from a real world network, such as a high clustering coefficient [29].
- **Scale-free networks** are characterized by a highly skewed distribution of contacts such that most of the nodes are weakly connected and a small number of nodes have very high connectivity [3]. There is empirical evidence from different research areas that in fact some real world networks exhibit such a skewed degree distribution [30, 23], varying over several orders of magnitude, although this is still debated for contact networks [2, 7]. The degree distribution of these heterogeneous networks can often approximated by a power-law behavior, which implies a non-negligible probability of finding nodes with very large degree.
- **Small world networks** are characterized by a degree distribution that is roughly symmetric about the mean, with a high degree of node clustering and a short characteristic path length [34]. This model, although better suited for social networks with high clustering coefficient, has a degree distribution and centrality measures decaying exponentially fast away from the average value. The small-world model thus generates homogeneous networks where the average of each metric is a typical value shared by all nodes of the network and with little variation.

However, the well-known mentioned families of networks do not cover all the aspects outlined for human interaction networks. In order to generate a more diverse set of networks we additionally use a **community-based graph generator** [14] that allows us to generate networks with all these parameters. Specifically, we perform a grid search of different values for the following parameters: i) Number of communities, ii) strength of those communities (probability of edges to be formed within communities), iii) the probability of also connecting to the neighbors of a node each nodes connects to, iv) number of communities a node can belong to, v) the probability of a node belonging to the multiple communities, vi) the strength of degree similarity effect on edge formation and vii) the strength of common neighbor’s effect on edge formation edges.

### 2.6 Network metrics computed

We have computed 30 different topological network metrics (i.e. that only use the adjacency matrix) for each of the network graphs. These can be categorised under:

- **Distance metrics** Wiener index, diameter, radius, local efficiency, global efficiency, eccentricity, closeness and betweenness. From these last 3 we compute the average, std and skewness.
- **Connection metrics** degree metrics (avg. degree, std. degree, max degree, min degree, branching factor, skewness degree, kurtosis degree, entropy degree), is connected, number of connected components, assortativity, clustering, transitivity and node connectivity.
- **Spectral metrics** algebraic connectivity and spectral radius.

We categorise these metrics in global and local metrics depending on whether they are computed only on the basis of nodes’ neighbours (local), or using the whole graph (global). Local metrics are all degree metrics, assortativity, clustering, local efficiency and transitivity. Global metrics are the remaining ones.

## 3 Experiments

We first show how networks with exactly the same amount of individuals and human interactions can lead to very different spread. All results use the same base simulator^1^ model and pathogen parameters. We then analyse all results on this highly diverse set of networks to study the networks metrics that influence the spread the most. Thus, the experiments and analysis in this paper can be divided into two differentiated parts: First, we simulate well-known families of networks with different assumptions about interactions between individuals and analyse the results in terms of the spread. Secondly, we compute a set of network metrics from the literature for each of these networks with the aim of finding the most predictive network metric for the spread of the epidemic. In this second part we also complement all the previous networks families with a network generator that is highly customisable, in order to generate an even larger set of networks, all with different network topology. The code and metrics for all network topologies will be made available at this project’s github page^2^.

### 3.1 Description of experiments

We have done experiments with the mentioned families of networks for different network sizes (N=500, 1000 and 2000) and levels of connectivity (mean degree of 4, 10 and 20). Previous work has found an average of 13.4 contacts per day per person consistently for different European countries [28]. However, people will make changes in behavior (e.g. reduce their number of contacts) in response to knowledge of an epidemic. These changes will not only reduce the number of contacts of the entire population, but also change the mixing patterns in the population, which is why we experiment with different average degree and types of human interactions.

Note that, independently of the mean degree of the population, all simulations have factored the same total of interactions per time step (i.e. so that on average each individual makes one contact per time step). This allows us to compare the results across different families of networks and levels of global/local interactions easily. Moreover, when we compare different families of networks we always compare results where the average degree is the same, but there is a different distribution of the edges of the contact network due to randomness and different network geometry.

For small-world networks the graph generator has a hyper-parameter that breaks clustering, increasing global connections (g). We experiment with different values of g (from 0.0 to 0.8, meaning that 80% of connections are random). Scale-free networks have clustering parameter (c), which we also experiment with. We run each simulation until the pathogen has naturally died off (i.e. herd immunity is in place). All the parameters for the pathogen has been set to those of covid-19. Specifically, *β* (transmission probability) is set to 0.155, *σ* to 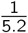 and *γ* to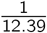. We report several statistics on the results of the simulations: i) percentage of nodes infected during the epidemic ii) peak of infected during the epidemic (also as a percentage of the total population) and iii) *R*_0_ (percentage of secondary infections per node infected in the first part of the simulation).

For each network family (e.g. Erdos Renyi) we have generated 30 random networks, and for each of these networks we have run the simulation 10 times. This is because there is both stochasticity in the generation of the graph as in the simulation itself. Additionally, after running the first simulation on the network (we denote this as the first generation) we have also reintroduced the pathogen and run the simulation on the remaining network in order to test whether herd immunity was in place (we name this the second generation).

### 3.2 Results with differently connected networks

We first analyse the spread over differently connected networks (average degree and levels of global and local connectivity) for an equal interaction budget. Again, this effectively means that for all the results presented, each individual makes one contact per time step, the difference in the results thus stems from the average degree of the population and the levels of local and global mixing. The results for different network sizes and connectivity can be seen in Table 1, where *N* represents the number of nodes or individuals in the network and *e* the average degree (edges). In this table we have aggregated all results for all network families (Erdos Renyi, Scale free and Small world) and we assume that each node has an equal interaction budget as presented in Section 3. As it was expected, networks with higher degree lead to a much higher percentage of infected before the pathogen naturally dies off and also a significantly higher peak of infection (compare for example the results for e=4 and e=20). However, the second generation is less pronounced (although if one sums up the infected from first and second generation for example for e=4 and e=20 the latter one still leads to much higher infection, i.e. 29% infected vs 74% for N=1000).

**Table 1:** Average ± standard deviation results for different network sizes (*n* nodes) and levels of connectivity (average *e* edges per node) assuming an equal budget of interaction per node (*B*_*i*_ = |*C*_*i*_ |). These results are aggregated over all three families of networks.

| Simulation with $p_g = 0.0$ and $p_l = 1.0$ | First generation | | | Second generation | | |
| --- | --- | --- | --- | --- | --- | --- |
| | Infected (%) | Peak (%) | $R_0$ | Infected (%) | Peak (%) | $R_0$ |
| e=4 N=500 | 27.2 ± 5.7 | 5.4 ± 0.6 | 1.3 ± 0.1 | 16.0 ± 2.1 | 4.5 ± 0.3 | 3.0 ± 0.5 |
| e=4 N=1000 | 17.9 ± 5.3 | 2.9 ± 0.4 | 1.3 ± 0.2 | 11.4 ± 2.3 | 2.5 ± 0.3 | 4.7 ± 1.1 |
| e=4 N=2000 | 11.7 ± 4.5 | 1.6 ± 0.3 | 1.4 ± 0.2 | 7.7 ± 2.1 | 1.4 ± 0.2 | 6.7 ± 2.1 |
| e=10 N=500 | 60.5 ± 9.3 | 8.7 ± 1.2 | 1.6 ± 0.2 | 11.0 ± 3.6 | 4.2 ± 0.3 | 1.7 ± 0.9 |
| e=10 N=1000 | 57.5 ± 11.9 | 6.6 ± 1.4 | 1.6 ± 0.2 | 7.2 ± 3.1 | 2.2 ± 0.3 | 2.6 ± 1.5 |
| e=10 N=2000 | 56.0 ± 13.8 | 5.5 ± 1.5 | 1.7 ± 0.2 | 4.4 ± 2.6 | 1.1 ± 0.2 | 3.4 ± 2.6 |
| e=20 N=500 | 70.8 ± 6.2 | 10.8 ± 1.2 | 1.7 ± 0.2 | 8.3 ± 3.3 | 4.1 ± 0.2 | 1.1 ± 0.8 |
| e=20 N=1000 | 68.9 ± 9.2 | 8.9 ± 1.5 | 1.7 ± 0.2 | 5.2 ± 3.6 | 2.1 ± 0.3 | 1.6 ± 1.8 |
| e=20 N=2000 | 67.7 ± 11.9 | 7.8 ± 1.7 | 1.8 ± 0.2 | 3.1 ± 2.8 | 1.1 ± 0.2 | 2.1 ± 2.8 |
| Simulation with $p_g = 0.1$ and $p_l = 0.9$ | First generation | | | Second generation | | |
| | Infected (%) | Peak (%) | $R_0$ | Infected (%) | Peak (%) | $R_0$ |
| e=4 N=500 | 40.5 ± 6.5 | 6.2 ± 0.7 | 1.4 ± 0.2 | 14.9 ± 2.5 | 4.4 ± 0.3 | 2.7 ± 0.6 |
| e=4 N=1000 | 33.3 ± 7.1 | 3.8 ± 0.6 | 1.4 ± 0.2 | 11.2 ± 2.6 | 2.4 ± 0.3 | 4.6 ± 1.3 |
| e=4 N=2000 | 28.2 ± 8.5 | 2.5 ± 0.5 | 1.4 ± 0.2 | 8.4 ± 2.6 | 1.4 ± 0.2 | 7.4 ± 2.6 |
| e=10 N=500 | 66.3 ± 5.0 | 9.5 ± 1.1 | 1.6 ± 0.2 | 9.1 ± 2.2 | 4.1 ± 0.1 | 1.3 ± 0.6 |
| e=10 N=1000 | 64.6 ± 5.8 | 7.6 ± 1.1 | 1.7 ± 0.2 | 5.5 ± 2.0 | 2.1 ± 0.1 | 1.8 ± 1.0 |
| e=10 N=2000 | 63.9 ± 6.3 | 6.6 ± 1.2 | 1.7 ± 0.2 | 3.2 ± 1.8 | 1.1 ± 0.1 | 2.2 ± 1.8 |
| e=20 N=500 | 73.4 ± 2.8 | 11.2 ± 1.0 | 1.7 ± 0.2 | 7.3 ± 1.2 | 4.0 ± 0.1 | 0.8 ± 0.3 |
| e=20 N=1000 | 72.5 ± 2.6 | 9.5 ± 1.1 | 1.8 ± 0.2 | 4.1 ± 1.0 | 2.0 ± 0.1 | 1.1 ± 0.5 |
| e=20 N=2000 | 71.9 ± 2.6 | 8.6 ± 1.1 | 1.8 ± 0.2 | 2.2 ± 0.6 | 1.0 ± 0.0 | 1.2 ± 0.6 |
| Simulation with $p_g = 0.2$ and $p_l = 0.8$ | First generation | | | Second generation | | |
| | Infected (%) | Peak (%) | $R_0$ | Infected (%) | Peak (%) | $R_0$ |
| e=4 N=500 | 52.5 ± 4.9 | 7.3 ± 0.7 | 1.4 ± 0.2 | 12.3 ± 2.3 | 4.2 ± 0.2 | 2.1 ± 0.58 |
| e=4 N=1000 | 49.2 ± 5.3 | 5.1 ± 0.6 | 1.5 ± 0.2 | 8.1 ± 2.2 | 2.2 ± 0.2 | 3.1 ± 1.11 |
| e=4 N=2000 | 47.3 ± 5.6 | 3.9 ± 0.6 | 1.5 ± 0.2 | 5.4 ± 2.2 | 1.2 ± 0.2 | 4.4 ± 2.20 |
| e=10 N=500 | 70.1 ± 2.9 | 10.3 ± 0.9 | 1.6 ± 0.2 | 8.0 ± 1.2 | 4.0 ± 0.1 | 1.0 ± 0.29 |
| e=10 N=1000 | 69.0 ± 2.6 | 8.6 ± 0.8 | 1.7 ± 0.2 | 4.6 ± 1.0 | 2.0 ± 0.1 | 1.3 ± 0.51 |
| e=10 N=2000 | 68.7 ± 2.3 | 7.6 ± 0.8 | 1.8 ± 0.2 | 2.4 ± 0.5 | 1.0 ± 0.0 | 1.4 ± 0.48 |
| e=20 N=500 | 74.9 ± 1.7 | 11.7 ± 0.8 | 1.7 ± 0.2 | 6.8 ± 0.6 | 4.0 ± 0.0 | 0.7 ± 0.15 |
| e=20 N=1000 | 74.1 ± 1.5 | 10.1 ± 0.8 | 1.8 ± 0.2 | 3.8 ± 0.4 | 2.0 ± 0.0 | 0.9 ± 0.21 |
| e=20 N=2000 | 73.6 ± 1.3 | 9.2 ± 0.7 | 1.9 ± 0.2 | 2.1 ± 0.5 | 1.0 ± 0.1 | 1.1 ± 0.51 |

Table 1 also includes results for different probabilities of local and global interaction (*p*_*l*_ and *p*_*g*_). Note that we always maintain *p*_*l*_ + *p*_*g*_ = 1 so that the same total number of human interactions is kept constant across the three simulations. The difference between the three cases is noteworthy. By decreasing the locality of how the pathogen spreads and allowing it to spread globally we can see that both the number of infections and the peak increase. Highly connected networks (e=20) increase their infection by around 4% when we compare the results of the *p*_*g*_ = 0.0 to *p*_*g*_ = 0.2. However, weakly connected networks (e=4) increase it by 25-30%. One can also appreciate the difference between decreasing the average degree of the population (e.g. going from e=20 to e=4) in a globally connected network (where there are random connections due to commuting, grocery shopping, etc.), where first generation infection decreases from 73.6 to 47.3% when N=2000, vs in a local network with *p*_*g*_ = 0.0, where it decreases from 67.7% to 11.7%.

#### Discussion of results

The first conclusion from our experiments is thus that not all human interactions affect the spread equally. In particular, higher levels of global connectivity increase the spread significantly, even when we are comparing the same amount of human interactions per time step. This is shown in our simulations in two ways: i) with networks of different average degree and ii) with simulations in which we vary *p*_*g*_. Thus, in a highly connected society, non-pharmaceutical interventions (such as social bubbles that may decrease the average degree for the population) may have less effect on decreasing the spread.

### 3.3 Results on well-known network families

We study now the effect of different well-known network families for equal and unequal interaction budgets. Let us begin with the equal interaction budget. In this case, at each time step, each individual contacts exactly one individual from their network (independently of the network size) with probability *p*_*l*_ and from the rest of the population with probability *p*_*g*_. The results for different network families can be seen in Table 1, where we have aggregated all results for the different network sizes and levels of connectivity.

The first striking observation is that Erdos Renyi generally leads to a higher percentage of infected than standard scale-free networks^3^. This is consistent with different results in the literature [19], which show that Erdos Renyi networks over-estimate the infection when compared to networks that resemble real-world ones better. This may be not intuitive at first, since scale-free networks represent networks with nodes with very high connectivity (e.g. super-spreaders). The hypothesis in the literature is that, assuming long lasting sterile immunity, highly connected nodes usually get infected quickly and then become immune, and given that they are highly connected they help slow down the spread [15]. It is noteworthy in any case that the infection progresses faster and a higher peak is achieved for scale-free networks, even when the total number of infected is lower. Increasing the clustering of scale-free networks (hyper-parameter c) decreases the spread even further as it is expected. This means that even in a network with a non-negligible percentage of highly connected individuals it helps to decrease global connections and increase the locality.

Small-world networks lead to the lowest infection rates. We observe, again, a more pronounced second generation of infections (still, total infection stays relatively low). However, the properties and structure of this network can be broken easily by increasing the amount of global connections (hyper-parameter q). A high percentage of random connections (q=0.8) leads to infection rates similar to those of Erdos Renyi networks. Once again, we can compare the results for different values of *p*_*l*_ and *p*_*g*_. In fact, increasing global interactions by setting *p*_*g*_ = 0.2 for scale-free and small world effectively means that all results get closer and closer to those of Erdos Renyi, and also that the herd immunity threshold increases (from 54% to 66% for Erdos Renyi).

We now compare these results to those with an unequal budget interaction, where highly connected individuals also have a larger number of interactions per time step, while the total of interactions for the population is maintained. The results are shown in Table 3. Interestingly, the same conclusions that we drawn from Table 2 still apply here: Erdos Renyi networks lead to larger spread than Scale-free, in this case by a larger margin. However, Scale-free networks lead to larger peak of infections. Small world networks lead to the least amount of infections. However, comparing each row in Table 2 to its corresponding row in Table 3 we can appreciate an increase in the infections when a heterogeneous budget of human interactions is assumed for the population. Note that in this paper we set a linear relationship between the budget of interactions and the degree of the individual. However, previous work has shown a super-linear relationship, meaning that the differences between Table 2 and 3 could be even more pronounced.

**Table 2:** Average ± standard deviation results for different network families assuming an equal budget of interaction per node. These results are aggregated over all network sizes and levels of connectivity.

| Simulation with $p_g = 0.0$ and $p_l = 1.0$ | First generation | | | Second generation | | |
| --- | --- | --- | --- | --- | --- | --- |
| | Infected | Peak | $R_0$ | Infected | Peak | $R_0$ |
| Erdos Renyi | 54.7 ± 20.7 | 7.0 ± 3.0 | 1.6 ± 0.2 | 7.1 ± 4.0 | 2.4 ± 1.3 | 2.4 ± 2.2 |
| Scale-free c=0.0 | 52.5 ± 22.1 | 7.1 ± 2.9 | 1.6 ± 0.3 | 7.7 ± 4.6 | 2.6 ± 1.3 | 2.7 ± 2.5 |
| Scale-free c=0.1 | 52.0 ± 22.6 | 7.2 ± 3.0 | 1.6 ± 0.3 | 7.8 ± 4.7 | 2.6 ± 1.3 | 2.8 ± 2.6 |
| Scale-free c=0.2 | 51.3 ± 22.8 | 7.1 ± 2.9 | 1.6 ± 0.3 | 7.7 ± 4.7 | 2.6 ± 1.3 | 2.7 ± 2.3 |
| Scale-free c=0.3 | 50.6 ± 23.5 | 7.0 ± 3.0 | 1.5 ± 0.3 | 7.9 ± 4.7 | 2.6 ± 1.3 | 2.9 ± 2.5 |
| Scale-free c=0.4 | 50.2 ± 23.4 | 7.0 ± 3.0 | 1.6 ± 0.3 | 7.9 ± 4.6 | 2.6 ± 1.3 | 2.8 ± 2.1 |
| Scale-free c=0.5 | 49.5 ± 24.0 | 6.9 ± 3.0 | 1.5 ± 0.3 | 7.9 ± 4.4 | 2.6 ± 1.3 | 2.8 ± 2.3 |
| Scale-free c=0.6 | 49.0 ± 24.0 | 6.8 ± 3.0 | 1.5 ± 0.3 | 7.9 ± 4.4 | 2.6 ± 1.3 | 2.8 ± 2.0 |
| Scale-free c=0.7 | 48.4 ± 24.9 | 6.8 ± 3.0 | 1.5 ± 0.3 | 7.9 ± 4.5 | 2.6 ± 1.3 | 2.7 ± 2.0 |
| Scale-free c=0.8 | 47.4 ± 24.7 | 6.6 ± 3.0 | 1.5 ± 0.3 | 7.9 ± 4.4 | 2.6 ± 1.4 | 2.8 ± 2.1 |
| Small-world q=0.0 | 19.8 ± 12.9 | 3.7 ± 2.0 | 1.5 ± 0.2 | 13.9 ± 6.0 | 3.0 ± 1.3 | 5.9 ± 3.4 |
| Small-world q=0.1 | 36.5 ± 21.0 | 4.5 ± 2.3 | 1.6 ± 0.2 | 11.0 ± 4.3 | 2.7 ± 1.3 | 4.5 ± 2.4 |
| Small-world q=0.2 | 44.5 ± 22.9 | 5.3 ± 2.5 | 1.6 ± 0.2 | 8.9 ± 4.0 | 2.5 ± 1.3 | 3.3 ± 1.6 |
| Small-world q=0.3 | 48.8 ± 23.2 | 5.9 ± 2.8 | 1.6 ± 0.2 | 8.1 ± 4.3 | 2.5 ± 1.3 | 2.8 ± 1.7 |
| Small-world q=0.4 | 50.9 ± 23.0 | 6.3 ± 2.9 | 1.6 ± 0.2 | 7.8 ± 4.3 | 2.5 ± 1.3 | 2.7 ± 2.0 |
| Small-world q=0.5 | 52.3 ± 22.7 | 6.7 ± 3.0 | 1.6 ± 0.2 | 7.5 ± 4.3 | 2.5 ± 1.3 | 2.6 ± 2.1 |
| Small-world q=0.6 | 53.4 ± 22.2 | 6.8 ± 3.0 | 1.6 ± 0.2 | 7.5 ± 4.5 | 2.5 ± 1.3 | 2.6 ± 2.1 |
| Small-world q=0.7 | 53.8 ± 22.0 | 6.9 ± 3.1 | 1.6 ± 0.2 | 7.4 ± 4.5 | 2.5 ± 1.3 | 2.6 ± 2.3 |
| Small-world q=0.8 | 54.0 ± 21.9 | 6.9 ± 3.0 | 1.6 ± 0.2 | 7.3 ± 4.3 | 2.5 ± 1.3 | 2.6 ± 2.5 |
| Simulation with $p_g = 0.1$ and $p_l = 0.9$ | First generation | | | Second generation | | |
| | Infected | Peak | $R_0$ | Infected | Peak | $R_0$ |
| Erdos Renyi | 60.9 ± 14.7 | 7.7 ± 2.7 | 1.6 ± 0.2 | 6.3 ± 3.5 | 2.4 ± 1.3 | 1.9 ± 1.6 |
| Scale-free c=0.0 | 58.9 ± 16.3 | 7.8 ± 2.7 | 1.6 ± 0.2 | 7.0 ± 4.2 | 2.5 ± 1.3 | 2.4 ± 2.3 |
| Scale-free c=0.1 | 59.0 ± 16.4 | 7.8 ± 2.7 | 1.6 ± 0.3 | 7.2 ± 4.6 | 2.5 ± 1.3 | 1.5 ± 2.5 |
| Scale-free c=0.2 | 58.4 ± 16.9 | 7.7 ± 2.7 | 1.6 ± 0.3 | 7.1 ± 4.3 | 2.5 ± 1.3 | 2.5 ± 2.4 |
| Scale-free c=0.3 | 57.8 ± 17.8 | 7.7 ± 2.8 | 1.6 ± 0.3 | 7.2 ± 4.5 | 2.5 ± 1.3 | 2.5 ± 2.7 |
| Scale-free c=0.4 | 57.0 ± 18.7 | 7.7 ± 2.9 | 1.6 ± 0.3 | 7.5 ± 4.6 | 2.5 ± 1.3 | 2.7 ± 3.1 |
| Scale-free c=0.5 | 56.6 ± 18.7 | 7.6 ± 2.8 | 1.6 ± 0.3 | 7.5 ± 4.6 | 2.5 ± 1.3 | 2.7 ± 2.9 |
| Scale-free c=0.6 | 56.0 ± 19.5 | 7.6 ± 2.9 | 1.6 ± 0.3 | 7.6 ± 4.7 | 2.6 ± 1.3 | 2.7 ± 2.8 |
| Scale-free c=0.7 | 55.0 ± 20.3 | 7.4 ± 2.9 | 1.6 ± 0.3 | 7.7 ± 4.6 | 2.6 ± 1.3 | 2.9 ± 2.9 |
| Scale-free c=0.8 | 54.3 ± 21.4 | 7.3 ± 3.0 | 1.6 ± 0.3 | 7.7 ± 4.9 | 2.6 ± 1.3 | 2.8 ± 2.9 |
| Small-world q=0.0 | 43.0 ± 19.2 | 4.9 ± 2.3 | 1.6 ± 0.2 | 10.7 ± 4.0 | 2.6 ± 1.3 | 4.3 ± 2.1 |
| Small-world q=0.1 | 50.4 ± 19.3 | 5.8 ± 2.4 | 1.6 ± 0.2 | 8.9 ± 4.2 | 2.5 ± 1.3 | 3.4 ± 2.3 |
| Small-world q=0.2 | 54.7 ± 18.4 | 6.4 ± 2.6 | 1.6 ± 0.2 | 7.9 ± 4.3 | 2.5 ± 1.3 | 2.8 ± 2.2 |
| Small-world q=0.3 | 57.2 ± 17.4 | 6.9 ± 2.8 | 1.6 ± 0.2 | 7.3 ± 4.1 | 2.4 ± 1.3 | 2.6 ± 2.3 |
| Small-world q=0.4 | 59.1 ± 15.9 | 7.2 ± 2.7 | 1.6 ± 0.2 | 6.9 ± 3.9 | 2.4 ± 1.3 | 2.3 ± 1.9 |
| Small-world q=0.5 | 60.2 ± 15.4 | 7.5 ± 2.8 | 1.6 ± 0.2 | 6.6 ± 3.8 | 2.4 ± 1.3 | 2.2 ± 1.9 |
| Small-world q=0.6 | 60.8 ± 15.0 | 7.6 ± 2.8 | 1.6 ± 0.2 | 6.4 ± 3.6 | 2.4 ± 1.3 | 2.0 ± 1.7 |
| Small-world q=0.7 | 61.2 ± 14.7 | 7.6 ± 2.8 | 1.6 ± 0.2 | 6.4 ± 3.6 | 2.4 ± 1.3 | 2.0 ± 1.7 |
| Small-world q=0.8 | 61.3 ± 14.6 | 7.7 ± 2.8 | 1.6 ± 0.2 | 6.3 ± 3.6 | 2.4 ± 1.3 | 2.0 ± 1.7 |
| Simulation with $p_g = 0.2$ and $p_l = 0.8$ | First generation | | | Second generation | | |
| | Infected | Peak | $R_0$ | Infected | Peak | $R_0$ |
| Erdos Renyi | 66.2 ± 9.8 | 8.5 ± 2.4 | 1.7 ± 0.2 | 5.3 ± 2.8 | 2.4 ± 1.3 | 1.4 ± 0.9 |
| Scale-free c=0.0 | 65.1 ± 10.5 | 8.5 ± 2.4 | 1.7 ± 0.2 | 5.8 ± 3.4 | 2.4 ± 1.3 | 1.6 ± 1.4 |
| Scale-free c=0.1 | 65.3 ± 10.3 | 8.6 ± 2.4 | 1.7 ± 0.2 | 5.7 ± 3.2 | 2.4 ± 1.3 | 1.6 ± 1.3 |
| Scale-free c=0.2 | 64.9 ± 10.7 | 8.5 ± 2.4 | 1.6 ± 0.2 | 5.8 ± 3.5 | 2.4 ± 1.3 | 1.7 ± 1.5 |
| Scale-free c=0.3 | 64.4 ± 11.3 | 8.5 ± 2.5 | 1.7 ± 0.2 | 6.0 ± 3.4 | 2.4 ± 1.3 | 1.8 ± 1.7 |
| Scale-free c=0.4 | 64.0 ± 11.9 | 8.5 ± 2.6 | 1.6 ± 0.3 | 6.1 ± 3.5 | 2.4 ± 1.3 | 2.0 ± 1.9 |
| Scale-free c=0.5 | 63.7 ± 11.8 | 8.4 ± 2.4 | 1.6 ± 0.2 | 6.2 ± 3.6 | 2.4 ± 1.3 | 1.9 ± 1.8 |
| Scale-free c=0.6 | 63.0 ± 12.7 | 8.3 ± 2.5 | 1.6 ± 0.3 | 6.4 ± 3.8 | 2.5 ± 1.3 | 2.0 ± 1.9 |
| Scale-free c=0.7 | 62.5 ± 13.4 | 8.2 ± 2.5 | 1.6 ± 0.2 | 6.5 ± 3.9 | 2.5 ± 1.3 | 2.1 ± 2.1 |
| Scale-free c=0.8 | 62.1 ± 13.6 | 8.1 ± 2.6 | 1.7 ± 0.3 | 6.6 ± 4.0 | 2.5 ± 1.3 | 2.2 ± 2.2 |
| Small-world q=0.0 | 58.4 ± 13.5 | 6.6 ± 2.3 | 1.7 ± 0.2 | 7.5 ± 3.6 | 2.4 ± 1.3 | 2.6 ± 1.7 |
| Small-world q=0.1 | 61.6 ± 12.1 | 7.2 ± 2.4 | 1.7 ± 0.2 | 6.7 ± 3.5 | 2.4 ± 1.3 | 2.1 ± 1.4 |
| Small-world q=0.2 | 63.4 ± 11.3 | 7.6 ± 2.4 | 1.7 ± 0.2 | 6.2 ± 3.3 | 2.4 ± 1.3 | 1.9 ± 1.3 |
| Small-world q=0.3 | 64.7 ± 10.4 | 8.0 ± 2.4 | 1.7 ± 0.2 | 5.8 ± 3.2 | 2.4 ± 1.3 | 1.7 ± 1.0 |
| Small-world q=0.4 | 65.6 ± 9.8 | 8.2 ± 2.4 | 1.7 ± 0.2 | 5.6 ± 3.0 | 2.4 ± 1.3 | 1.6 ± 0.9 |
| Small-world q=0.5 | 66.2 ± 9.3 | 8.3 ± 2.4 | 1.7 ± 0.2 | 5.4 ± 2.8 | 2.4 ± 1.3 | 1.5 ± 1.0 |
| Small-world q=0.6 | 66.6 ± 9.1 | 8.4 ± 2.4 | 1.7 ± 0.2 | 5.3 ± 2.8 | 2.4 ± 1.3 | 1.4 ± 0.8 |
| Small-world q=0.7 | 66.6 ± 9.3 | 8.5 ± 2.4 | 1.7 ± 0.2 | 5.3 ± 2.9 | 2.4 ± 1.3 | 1.4 ± 0.8 |
| Small-world q=0.8 | 66.6 ± 9.3 | 8.5 ± 2.4 | 1.7 ± 0.2 | 5.3 ± 2.9 | 2.4 ± 1.3 | 1.4 ± 0.9 |

**Table 3:** Average ± standard deviation results for different network families assuming a budget of interaction dependent on the degree of each node. These results are aggregated over all network sizes and levels of connectivity.

| Simulation with $p_g = 0.0$ and $p_l = 1.0$ | First generation | | | Second generation | | |
| --- | --- | --- | --- | --- | --- | --- |
| | Infected | Peak | $R_0$ | Infected | Peak | $R_0$ |
| Erdos Renyi | 61.6 ± 11.6 | 8.3 ± 2.4 | 1.7 ± 0.2 | 5.1 ± 2.6 | 2.4 ± 1.3 | 1.3 ± 0.8 |
| Scale-free c=0.0 | 60.4 ± 7.3 | 12.2 ± 1.8 | 2.3 ± 0.4 | 3.8 ± 2.0 | 2.3 ± 1.3 | 0.6 ± 0.2 |
| Scale-free c=0.1 | 60.3 ± 7.4 | 12.3 ± 1.9 | 2.4 ± 0.4 | 3.8 ± 2.0 | 2.3 ± 1.3 | 0.7 ± 0.3 |
| Scale-free c=0.2 | 59.9 ± 7.6 | 12.4 ± 1.9 | 2.4 ± 0.5 | 3.8 ± 2.0 | 2.3 ± 1.3 | 0.7 ± 0.3 |
| Scale-free c=0.3 | 59.6 ± 7.8 | 12.4 ± 2.0 | 2.5 ± 0.5 | 3.8 ± 2.0 | 2.3 ± 1.3 | 0.7 ± 0.3 |
| Scale-free c=0.4 | 59.1 ± 8.3 | 12.4 ± 2.1 | 2.6 ± 0.6 | 3.9 ± 2.1 | 2.3 ± 1.3 | 0.7 ± 0.4 |
| Scale-free c=0.5 | 59.0 ± 8.6 | 12.4 ± 2.2 | 2.6 ± 0.5 | 3.9 ± 2.1 | 2.4 ± 1.3 | 0.7 ± 0.4 |
| Scale-free c=0.6 | 58.8 ± 9 | 12.3 ± 2.2 | 2.6 ± 0.5 | 4.0 ± 2.1 | 2.4 ± 1.3 | 0.8 ± 0.4 |
| Scale-free c=0.7 | 58.4 ± 9.4 | 12.2 ± 2.1 | 2.6 ± 0.5 | 4.1 ± 2.2 | 2.4 ± 1.3 | 0.8 ± 0.5 |
| Scale-free c=0.8 | 57.8 ± 10.2 | 11.9 ± 2.2 | 2.5 ± 0.5 | 4.3 ± 2.3 | 2.4 ± 1.3 | 0.9 ± 0.7 |
| Small-world q=0.0 | 20.2 ± 13.3 | 3.7 ± 2.0 | 1.5 ± 0.2 | 14.1 ± 6.0 | 3.1 ± 1.4 | 6.0 ± 3.5 |
| Small-world q=0.1 | 37.0 ± 21.3 | 4.6 ± 2.3 | 1.6 ± 0.2 | 11.0 ± 4.3 | 2.7 ± 1.3 | 4.5 ± 2.4 |
| Small-world q=0.2 | 45.8 ± 22.5 | 5.5 ± 2.6 | 1.6 ± 0.2 | 8.8 ± 4.1 | 2.5 ± 1.3 | 3.3 ± 1.7 |
| Small-world q=0.3 | 50.2 ± 22.1 | 6.2 ± 2.8 | 1.6 ± 0.2 | 8.0 ± 4.2 | 2.5 ± 1.3 | 2.9 ± 2.0 |
| Small-world q=0.4 | 53.0 ± 20.9 | 6.6 ± 2.8 | 1.6 ± 0.2 | 7.5 ± 4.3 | 2.5 ± 1.3 | 2.7 ± 2.3 |
| Small-world q=0.5 | 55.2 ± 19.4 | 7 ± 2.9 | 1.6 ± 0.2 | 7.1 ± 4.1 | 2.4 ± 1.3 | 2.4 ± 2.2 |
| Small-world q=0.6 | 56.2 ± 18.4 | 7.2 ± 2.9 | 1.6 ± 0.2 | 6.9 ± 4.0 | 2.4 ± 1.3 | 2.4 ± 2.1 |
| Small-world q=0.7 | 57.2 ± 17.5 | 7.4 ± 2.9 | 1.6 ± 0.2 | 6.7 ± 3.9 | 2.4 ± 1.3 | 2.2 ± 1.9 |
| Small-world q=0.8 | 57.5 ± 17.5 | 7.4 ± 2.9 | 1.6 ± 0.2 | 6.7 ± 3.9 | 2.4 ± 1.3 | 2.2 ± 2.1 |
| Simulation with $p_g = 0.1$ and $p_l = 0.9$ | First generation | | | Second generation | | |
| | Infected | Peak | $R_0$ | Infected | Peak | $R_0$ |
| Erdos Renyi | 65.0 ± 8.8 | 9.0 ± 2.0 | 1.7 ± 0.2 | 4.7 ± 2.4 | 2.4 ± 1.3 | 1.1 ± 0.4 |
| Scale-free c=0.0 | 61.8 ± 5.8 | 12.2 ± 1.5 | 2.3 ± 0.4 | 3.7 ± 1.9 | 2.3 ± 1.3 | 0.6 ± 0.2 |
| Scale-free c=0.1 | 61.5 ± 6 | 12.3 ± 1.6 | 2.4 ± 0.4 | 3.7 ± 1.9 | 2.3 ± 1.3 | 0.6 ± 0.2 |
| Scale-free c=0.2 | 61.2 ± 6.1 | 12.4 ± 1.7 | 2.4 ± 0.4 | 3.8 ± 2.0 | 2.3 ± 1.3 | 0.6 ± 0.4 |
| Scale-free c=0.3 | 61.2 ± 6.1 | 12.5 ± 1.7 | 2.5 ± 0.4 | 3.7 ± 1.9 | 2.3 ± 1.3 | 0.6 ± 0.2 |
| Scale-free c=0.4 | 60.5 ± 6.5 | 12.4 ± 1.8 | 2.5 ± 0.5 | 3.7 ± 1.9 | 2.3 ± 1.3 | 0.6 ± 0.3 |
| Scale-free c=0.5 | 60.5 ± 6.7 | 12.3 ± 1.9 | 2.5 ± 0.5 | 3.8 ± 2.0 | 2.3 ± 1.3 | 0.7 ± 0.3 |
| Scale-free c=0.6 | 60.4 ± 6.9 | 12.2 ± 1.8 | 2.5 ± 0.5 | 3.8 ± 2.0 | 2.3 ± 1.3 | 0.7 ± 0.3 |
| Scale-free c=0.7 | 60.3 ± 7.1 | 12.1 ± 1.8 | 2.5 ± 0.5 | 3.9 ± 2.0 | 2.3 ± 1.3 | 0.7 ± 0.3 |
| Scale-free c=0.8 | 60.2 ± 7.5 | 12 ± 1.9 | 2.5 ± 0.5 | 3.9 ± 2.0 | 2.3 ± 1.3 | 0.7 ± 0.3 |
| Small-world q=0.0 | 43.3 ± 19.3 | 4.9 ± 2.3 | 1.6 ± 0.2 | 10.8 ± 4.0 | 2.6 ± 1.3 | 4.4 ± 2.1 |
| Small-world q=0.1 | 51.5 ± 19 | 5.9 ± 2.5 | 1.6 ± 0.2 | 8.6 ± 4.2 | 2.5 ± 1.3 | 2.2 ± 2.2 |
| Small-world q=0.2 | 56.1 ± 16.8 | 6.6 ± 2.6 | 1.6 ± 0.2 | 7.5 ± 3.9 | 2.5 ± 1.3 | 2.7 ± 2.1 |
| Small-world q=0.3 | 59.1 ± 14.7 | 7.2 ± 2.6 | 1.7 ± 0.2 | 6.7 ± 3.6 | 2.4 ± 1.3 | 2.2 ± 1.8 |
| Small-world q=0.4 | 61.1 ± 13.3 | 7.6 ± 2.6 | 1.7 ± 0.2 | 6.1 ± 3.2 | 2.4 ± 1.3 | 1.9 ± 1.4 |
| Small-world q=0.5 | 62.3 ± 12.4 | 7.9 ± 2.6 | 1.7 ± 0.2 | 5.8 ± 3.1 | 2.4 ± 1.3 | 1.7 ± 1.2 |
| Small-world q=0.6 | 63.0 ± 11.4 | 8.1 ± 2.5 | 1.7 ± 0.2 | 5.6 ± 3.0 | 2.4 ± 1.3 | 1.6 ± 1.0 |
| Small-world q=0.7 | 63.5 ± 11.4 | 8.2 ± 2.5 | 1.7 ± 0.2 | 5.4 ± 2.8 | 2.4 ± 1.3 | 1.5 ± 1.0 |
| Small-world q=0.8 | 63.4 ± 11.1 | 8.3 ± 2.5 | 1.7 ± 0.2 | 5.4 ± 3.0 | 2.4 ± 1.3 | 1.5 ± 0.9 |
| Simulation with $p_g = 0.2$ and $p_l = 0.8$ | First generation | | | Second generation | | |
| | Infected | Peak | $R_0$ | Infected | Peak | $R_0$ |
| Erdos Renyi | 67.4 ± 6.8 | 9.6 ± 1.7 | 1.8 ± 0.2 | 4.3 ± 2.1 | 2.3 ± 1.3 | 0.9 ± 0.3 |
| Scale-free c=0.0 | 62.7 ± 5.0 | 12.1 ± 1.3 | 2.3 ± 0.4 | 3.7 ± 1.9 | 2.3 ± 1.3 | 0.6 ± 0.2 |
| Scale-free c=0.1 | 62.4 ± 5 | 12.2 ± 1.4 | 2.3 ± 0.3 | 3.7 ± 1.9 | 2.3 ± 1.3 | 0.6 ± 0.2 |
| Scale-free c=0.2 | 62.2 ± 5.1 | 12.2 ± 1.4 | 2.3 ± 0.4 | 3.7 ± 1.9 | 2.3 ± 1.3 | 0.6 ± 0.2 |
| Scale-free c=0.3 | 62.0 ± 5.1 | 12.3 ± 1.5 | 2.4 ± 0.4 | 3.7 ± 1.9 | 2.3 ± 1.3 | 0.6 ± 0.2 |
| Scale-free c=0.4 | 61.8 ± 5.3 | 12.3 ± 1.6 | 2.5 ± 0.5 | 3.7 ± 1.9 | 2.3 ± 1.3 | 0.6 ± 0.2 |
| Scale-free c=0.5 | 61.8 ± 5.2 | 12.3 ± 1.6 | 2.5 ± 0.5 | 3.7 ± 1.9 | 2.3 ± 1.3 | 0.6 ± 0.2 |
| Scale-free c=0.6 | 61.7 ± 5.6 | 12.2 ± 1.5 | 2.5 ± 0.4 | 3.7 ± 1.9 | 2.3 ± 1.3 | 0.6 ± 0.2 |
| Scale-free c=0.7 | 61.5 ± 5.6 | 12.1 ± 1.5 | 2.5 ± 0.5 | 3.8 ± 1.9 | 2.3 ± 1.3 | 0.6 ± 0.2 |
| Scale-free c=0.8 | 61.5 ± 5.8 | 12 ± 1.5 | 2.4 ± 0.5 | 3.8 ± 1.9 | 2.3 ± 1.3 | 0.7 ± 0.2 |
| Small-world q=0.0 | 58.0 ± 13.4 | 6.5 ± 2.3 | 1.6 ± 0.2 | 7.7 ± 3.7 | 2.5 ± 1.3 | 2.7 ± 1.7 |
| Small-world q=0.1 | 61.7 ± 11.7 | 7.3 ± 2.4 | 1.7 ± 0.2 | 6.6 ± 3.5 | 2.4 ± 1.3 | 2.0 ± 1.2 |
| Small-world q=0.2 | 64.2 ± 9.9 | 7.8 ± 2.3 | 1.7 ± 0.2 | 5.8 ± 3.0 | 2.4 ± 1.3 | 1.7 ± 0.9 |
| Small-world q=0.3 | 65.2 ± 9.1 | 8.2 ± 2.2 | 1.7 ± 0.2 | 5.5 ± 2.8 | 2.4 ± 1.3 | 1.5 ± 0.9 |
| Small-world q=0.4 | 66.3 ± 8.4 | 8.5 ± 2.2 | 1.7 ± 0.2 | 5.2 ± 2.7 | 2.4 ± 1.3 | 1.3 ± 0.7 |
| Small-world q=0.5 | 67.1 ± 7.8 | 8.8 ± 2.1 | 1.7 ± 0.2 | 4.9 ± 2.4 | 2.4 ± 1.3 | 1.2 ± 0.6 |
| Small-world q=0.6 | 67.1 ± 7.8 | 8.8 ± 2.1 | 1.7 ± 0.2 | 4.9 ± 2.4 | 2.4 ± 1.3 | 1.2 ± 0.5 |
| Small-world q=0.7 | 67.3 ± 7.7 | 9 ± 2.1 | 1.7 ± 0.2 | 4.8 ± 2.4 | 2.4 ± 1.3 | 1.1 ± 0.5 |
| Small-world q=0.8 | 67.5 ± 7.5 | 9 ± 2.1 | 1.7 ± 0.2 | 4.8 ± 2.4 | 2.4 ± 1.3 | 1.1 ± 0.5 |

#### Discussion of results

Erdos Renyi networks, the assumption of most epidemiological models, often over-estimate the infection when compared to networks that resemble real-world ones better. Powerlaw often show less infections than Erdos Renyi but a higher peak. Small world networks are consistently resilient to pathogen spread. However, slightly increasing the globality of small world networks while keeping the same number of interactions leads to a significantly higher infection rate. A heterogeneous budget of human interactions leads to an increased number of infections compared to a homogeneous one.

### 3.4 Influence of network metrics on the spread

We now put together all the contact networks that we have generated and used in our simulations. This includes not only all the well-known families of networks that we analysed in our previous section but also the community-based networks. The total of networks analysed in this experiment is 9061. We thus build a large dataset of social networks. For each one of these networks we have computed the 30 network metrics mentioned before, together with some statistics of the spread, i.e. average percentage of individuals infected and peak of infection across 10 simulations. We analyse how different network metrics influence the spread of the pathogen for a large range of network topologies.

### 3.4.1 Univariate analysis

A correlation analysis can be found in Table 4. The conclusions are as follows:

**Table 4:** Average ± standard deviation results for different network families assuming an equal budget of interaction per node. These results are aggregated over all network sizes and levels of connectivity.

|  | Equal distr. of interactions |  |  |  | Unequal distr. of interactions |  |  |  |
| --- | --- | --- | --- | --- | --- | --- | --- | --- |
|  | Pearson corr. |  | Spearman rank corr. |  | Pearson corr. |  | Spearman rank corr. |  |
| Network metrics | Infected | Peak | Infected | Peak | Infected | Peak | Infected | Peak |
| algebraic conn. | 0.57 | 0.57 | 0.66 | 0.45 | 0.66 | 0.63 | 0.17 | -0.06 |
| assort. corr. | 0.08 | -0.06 | -0.06 | -0.07 | -0.15 | -0.14 | 0.12 | -0.13 |
| avg betweenness | -0.32 | -0.39 | -0.22 | -0.28 | -0.41 | -0.35 | -0.34 | -0.32 |
| avg closeness | 0.82 | 0.87 | 0.83 | 0.87 | 0.85 | 0.81 | 0.74 | 0.65 |
| avg degree | 0.76 | 0.82 | 0.75 | 0.75 | 0.69 | 0.73 | 0.36 | 0.40 |
| avg eccentricity | -0.33 | -0.92 | -0.27 | -0.89 | -0.45 | -0.90 | -0.32 | -0.66 |
| branching factor | 0.15 | 0.08 | 0.37 | 0.19 | 0.23 | -0.58 | 0.39 | -0.56 |
| clustering | -0.34 | -0.39 | -0.31 | -0.31 | -0.36 | -0.40 | -0.10 | 0.06 |
| entropy degree | 0.49 | 0.20 | 0.41 | 0.27 | 0.32 | 0.08 | 0.75 | 0.57 |
| is connected | -0.07 | 0.14 | 0.02 | 0.05 | 0.14 | 0.23 | -0.33 | -0.29 |
| kurtosis degree | -0.34 | -0.09 | -0.26 | 0.01 | -0.06 | -0.09 | 0.14 | 0.66 |
| local eff. | -0.16 | -0.23 | -0.14 | -0.16 | -0.26 | -0.28 | 0.07 | 0.18 |
| node conn. | 0.38 | 0.47 | 0.41 | 0.33 | 0.48 | 0.53 | -0.16 | -0.22 |
| skew betweenness | -0.18 | -0.14 | -0.11 | -0.03 | 0.00 | -0.14 | 0.44 | 0.65 |
| skew closeness | 0.23 | 0.26 | 0.32 | 0.23 | 0.32 | 0.32 | 0.31 | 0.21 |
| skew degree | -0.22 | -0.09 | -0.13 | 0.02 | 0.00 | -0.08 | 0.42 | 0.67 |
| skew eccentricity | 0.01 | 0.02 | 0.03 | 0.01 | 0.01 | -0.03 | 0.00 | 0.08 |
| spectral radius | 0.55 | 0.32 | 0.51 | 0.42 | 0.35 | 0.15 | 0.80 | 0.79 |
| std betweenness | -0.30 | -0.26 | -0.14 | -0.10 | -0.09 | -0.23 | 0.32 | 0.54 |
| std closeness | 0.10 | -0.15 | 0.09 | 0.03 | -0.04 | -0.25 | 0.73 | 0.66 |
| std degree | 0.37 | 0.26 | 0.38 | 0.37 | 0.21 | 0.13 | 0.82 | 0.91 |
| std eccentricity | -0.45 | -0.54 | -0.42 | -0.44 | -0.42 | -0.54 | 0.03 | 0.04 |
| transitivity | -0.14 | -0.18 | -0.19 | -0.16 | -0.31 | -0.26 | -0.30 | -0.05 |
| global eff. | 0.84 | 0.87 | 0.85 | 0.87 | 0.87 | 0.81 | 0.75 | 0.66 |
| diameter | -0.34 | -0.89 | -0.27 | -0.84 | -0.46 | -0.90 | -0.32 | -0.55 |
| radius | -0.32 | -0.80 | -0.26 | -0.80 | -0.44 | -0.79 | -0.32 | -0.76 |

- Global efficiency is the most predictive metric overall (not only of total of infected but also of the peak). The relationship seems to be linear, as indicated by the Pearson correlation. Average closeness follows. Both of these metrics are novel to the literature.
- Metrics such as the spectral radius are not as predictive of infections as was suggested in the literature (at least not on their own). The same is applicable to clustering and transitivity.
- Some metrics seem to be predictive of the infected, but not of the peak. This is the case of the average degree, and more specifically for a unequal budget of interactions.
- There are some metrics that are not linearly correlated but show great correlation in rank, for example diameter, radius and avg eccentricity, with up to 0.92 negative correlation.
- The skewness of the degree distribution (related to super spreaders) alone is not enough for predicting the spread.

We display in Figure 1.1.3 some of these network metrics (x axis) versus the percentage of infected (y axis). The first row shows some metrics of the degree distribution, which alone do not seem sufficient to predict infections. The next row (spectral radius and clustering) shows metrics that have been proposed in the literature as predictive of the spread. We can see that these metrics, alone, also do not show a clear relationship with infections. Finally, the last row shows metrics that are novel (global efficiency and algebraic connectivity) which have a clear influence on the spread.

#### Discussion of results

The correlation analysis shows that global efficiency and average closeness are the most predictive network metrics. This contradicts previous studies which suggested average of the degree distribution, clustering and spectral radius to be most informative. Studies aimed at measuring the average degree across the population exist, however we show that even for a fixed average degree the spread can change drastically (i.e. from 10% to 50% infection for average degree of 4, and from 30% to more than 80% for average degree of 20).

### 3.4.2 Multivariate analysis

We investigated the influence of metrics on the pathogen spread with a multivariate linear regression analysis. The results are shown in Figure 1.1.3. We report adjusted *R*^2^ computed on the test set of 100 random holdouts with 80% of data for training and 20% for testing. *R*^2^ is a goodness-of-fit measure for linear regression models that indicates the percentage of the variance in the dependent variable that the independent variables explain collectively. The *R*^2^ obtained using all the network metrics is 0.934. This value indicates that the independent variables help us explain the spread almost perfectly. The remaining variance of the data could be partly due to the randomness of the simulation. This figure also shows the predictive power of different groups of metrics. First, we see that average degree alone is not enough to predict the spread accurately. Considering additional degree metrics (skewness, entropy, etc.) improves the predictive performance (*R*^2^ from 0.473 to 0.631), further improved when taking into account the remaining local metrics (*R*^2^ from 0.631 to 0.710). However, the model built using only global efficiency achieves better performance than the model that uses all degree metrics and the model that uses all local metrics (*R*^2^ of 0.747). Additional global metrics complement global efficiency and increase the performance further. A non-linear regression model trained with global metrics alone was able to accurately predict the spread as well as a model trained with all metrics.

However, there is a very strong multicollinearity (indicated by the condition number) in these models, which makes the interpretation of the models not reliable, which is why we do not interpret the linear regression coefficients. Multicollinearity is a common problem when estimating linear linear models. It occurs when there are high correlations among predictor variables, leading to unreliable and unstable estimates of regression coefficients. In order to reduce multicollinearity and be able to interpret the final regression model we perform a exhaustive feature selection that tests a large amount of combinations of features (all sets from 3 to 12 features) using 3 fold cross-validation. We observe that many different sets of features still explain the data variance greatly (with an *R*^2^ of 0.837 for only 3 selected features) and reject that multicolinearity was present. This indicates that there are combinations of these features that can reliably explain the variance in the spread.

#### Discussion of results

A combination of network metrics can accurately predict the variability on the spread for both equal and unequal budget of human interactions. Linear regression analysis shows that global efficiency has the highest influence on the spread. This metric relate to the path lengths between nodes and global connectivity of the social graph. Both contact tracing or mobility data that are already being recorded can be used for example to infer path lengths in different societies.

## 4 Conclusions

Among other research findings, our simulations show i) how much the topology of social networks can highly influence pathogen spread, ii) that reducing incidental casual contact reduces significantly the spread, iii) that small world network topologies are resilient to infections, iv) the usefulness of a principled multivariate approach for evaluating metrics predictive of the spread and v) the superiority of metrics that relate to the graph path length over metrics of the degree distribution. Our simulations also highlight the importance of modeling a wide range of parameters of a social network (and not only the average but also the distribution), for example the degree and contact network per individual, mobility, connectivity, etc. This could be done either through behavioural studies, contact tracing or mobility data that is already being recorded. Measuring these for a population would lead to much more accurate epidemiological simulations and social interventions. Our results also show that the effect of a social intervention depends on the underlying social network. This is of crucial importance. A social intervention will almost always vary in its effect on each property of a social network. For example, closing public spaces reduces long range connectivity but not necessarily short range connectivity. Social interventions can be then seen as transitions from one complex social network organisation to another, not merely attenuations or amplifications of any given network. As different previous works have already pointed out, to maximise the impact of social interventions, we need complex, prescriptive models of social networks.

The space of optimal interventions could be large, diverse, and non-intuitive: the same intervention can have radically different (indeed opposing effects) across different real-world networks, and the same outcome may require radically different interventions to achieve. Given the observed wide space of scenarios, interventional inference is likely to require an exploration of the graph properties of the networks scenarios assessed to derive maximal efficient change in those properties that leads to the desired optimal outcome. It is thus clear that we need a wide range of complex network analysis epidemiological tools, as well as a platform that allows to do so systematically. Even if we do not know what are the topologies of real-world networks (and this certainly can change across societies and regions), it is worth exploring the space of complex interactions scenarios in a systematic manner. This could lead to a subset of interventions that perhaps collapse in trend regardless of the network and help restrict the decision making space.

Contact-tracing data specifically provide information to reconstruct transmission chains and understand outbreak dynamics [32, 16]. These data can in turn generate valuable intelligence on key epidemiological parameters and risk factors for transmission [27], which paves the way for more-targeted and cost-effective interventions. Determining a complete mixing network requires knowledge of every individual in a population and every relationship between individuals. For all but the smallest populations, this is an impractically time-consuming task. However, since long lasting sterile immunity may not be guaranteed, this may be crucial to understand how different behavioural factors and social networks influence the spread. Specifically, these network metrics can be captured from a subgraph of the population instead of the whole network. They may be useful to design more accurate simulations and to recommend interventions or to target/correct aspects of connectivity that will actually control infection spread, while maintaining economic activity. Among the studied families of social graphs, small world networks excel in their resilience to infections.

We believe that further study into the effect of different social interventions on the spread and global efficiency of the network are needed. We think that such a line of research could potentially lead to crafting measures that define the optimal interventions in terms of spread per social contact. A promising metric could be the participation coefficient [11], which could be extracted from contact tracing, and would give an indication of how “provincial” each node in the graph is. Interventions aimed at the most connected nodes (e.g. vaccination) could potentially decrease global efficiency to a large extent.

## Data Availability

The data is generated via simulations. The code and dataset will be made available at https://github.com/mperezortiz/topology_spread

## Footnotes

1 https://github.com/ryansmcgee/seirsplus

2 https://github.com/mperezortiz/topology_spread

3 Here we refer to scale-free with c=0.0 and small-world with q=0.0, as these are additional hyper-parameters of the graph generators that change the network topology.

